# Measuring the Latent Tuberculosis Infection Care Cascade Using Electronic Health Record Data from Primary Care Clinics in the Tuberculosis Epidemiologic Studies Consortium-III

**DOI:** 10.1101/2025.08.25.25333908

**Authors:** Laura A. Vonnahme, Preeti Ravindhran, Julie Espey, Bhumika Sharma, Taylor Moore, Kaylynn Aiona, Jacek Skarbinski, Masahiro Narita, Priya B. Shete, Jagadheeswari Adhimurthy, Richard Broadhurst, Paul Wada, Grace Bond, Matthew T. Murrill, Kuan-Chieh Huang, Meagan Lee, Jihming Lin, Kathryn Winglee, the Tuberculosis Epidemiologic Studies Consortium

## Abstract

**Importance:** Tuberculosis (TB) was the leading infectious cause of death worldwide in 2023. Most US TB cases represent reactivation of latent TB infection (LTBI). Because LTBI treatment is approximately 90% effective for preventing TB disease, LTBI screening and treatment are primary strategies for US TB elimination. The Tuberculosis Epidemiologic Studies Consortium-III (TBESC-III) supports US TB elimination efforts by addressing LTBI among individuals at higher risk of infection seeking care in US primary care clinics.

**Objective:** To characterize and measure outcomes across a longitudinal LTBI care cascade, from proportion of persons at higher risk through testing, diagnosis, and treatment.

**Design:** Longitudinal study using patient-level electronic health record (EHR) data.

**Setting:** Primary care clinics serving at least 10,000 non–US-born individuals annually from countries with high TB disease incidence rates (defined as ≥10 cases per 100,000 persons among expatriates living in the US).

**Participants:** Persons at higher risk of TB infection, defined as non-US birth or, if unknown, a non-English language preference, who had a visit during the study period at a participating clinic.

**Intervention(s) (for clinical trials) or Exposure(s) (for observational studies):** Not relevant

**Main Outcome(s) and Measure(s):** Among participants without prior TB or LTBI testing, diagnosis, or treatment documented (ie, cascade eligible), we determined proportions: tested for TB infection, with available test results, with positive results, chest imaging ordered, LTBI diagnoses, and with LTBI treatment prescribed, started, and completed. Percentages were averaged across four sites representing multiple clinics.

**Results:** Of 3.5 million persons seeking care, on average, 48% were at higher risk of TB infection, and 69% of these were cascade eligible. Among cascade eligible individuals, 14% were tested; 92% of those tested had available results, with 17% testing positive. Of those testing positive, 82% underwent chest imaging; 70% met LTBI diagnostic criteria. Among those diagnosed, 61% were prescribed treatment; 87% started treatment, with 56% completing treatment.

**Conclusions and Relevance:** Although an average of 17% of participants tested had TB infection, most (average, 86%) higher-risk individuals were not tested; an average of 39% of those diagnosed were not prescribed treatment, and nearly half (average, 44%) did not complete treatment. Targeted interventions to increase LTBI testing and treatment completion among higher-risk individuals could facilitate more preventive treatment and reductions in TB-associated morbidity.

## Introduction

Tuberculosis (TB) elimination in the US is defined as a national incidence rate of <1 case per 1,000,000 population. Currently, the incidence rate is 27 times the elimination threshold.^1,2^ The majority of TB disease in the US is due to reactivation of latent TB infection (LTBI), disproportionately among non–US-born persons likely infected in their countries of birth, despite LTBI treatment being approximately 90% effective for preventing TB disease progression.^3–9^

The Centers for Disease Control and Prevention (CDC) and the US Preventive Services Task Force (USPSTF) recommend TB screening for persons who were born in or have lived outside the US in countries where TB is more prevalent.^10–12^ However, TB screening among individuals at higher risk in the United States is inconsistently implemented, and LTBI treatment completion rates are historically low.^7,13–16^ Increasing LTBI treatment among non–US-born persons in the US might be one of the most effective tools for TB elimination.^17^ Although most TB care is performed in public health clinics, primary care clinics represent a critical setting for this scale-up, as many provide routine medical care for persons at higher risk of infection.^13,14,18–23^

The LTBI care cascade defines steps from identifying individuals at increased risk for TB infection through to treatment completion. Assessing patients eligible for and receiving care at each step is a useful for identifying gaps across the cascade, which can be used to improve TB prevention in health care systems.^15,24,25^ However, defining and evaluating the LTBI care cascade using electronic health record (EHR) data from primary care settings poses several challenges.^15,26,27^ EHR data are not collected for the purpose of developing care cascades; key risk factors are often missing or not standardized, and there are no variables to indicate definitive LTBI diagnosis or treatment outcomes.^14,28,29^ EHR data elements are not standardized across health care systems or even individual clinics. Furthermore, the longitudinal nature of the care cascade, including developing standardized time allowed between steps, has not been previously considered.

The third iteration of the Tuberculosis Epidemiologic Studies Consortium (TBESC-III) aims to improve LTBI care cascade outcomes among non–US-born populations at higher risk of infection who seek care in primary care settings.^30^ The objective of this analysis was to standardize the methodology for defining the LTBI care cascade and assess outcomes in TBESC-III primary care networks using longitudinal EHR data to guide future interventions to improve LTBI care. This care cascade provides a baseline understanding of LTBI testing and treatment outcomes in primary care clinics, identifying specific steps for proposing interventions to improve outcomes across the LTBI care cascade.

## Methods

### Study Design and Population

Detailed information on TBESC-III consortium study design and population is available (eMethods 1 in **Supplement 1**). Briefly, the consortium has 4 sites; all primary care health care systems serving at least 10,000 non–US-born individuals annually from countries with high TB disease incidence rates (defined as ≥10 cases per 100,000 persons among expatriates living in the US).^31^ Study participants had to have (1) reported non-US birth or, if unknown, a non-English language preference, (2) a clinic visit during the study period at a participating clinic and (3) meet additional site-specific eligibility criteria (Table 1). EHR data were extracted on study participants’ demographics, visits, and imaging that occurred during the study period and within the 2 years preceding it. All available EHR data on TB- and LTBI-related diagnostic testing; *International Classification of Diseases*, *Ninth Revision* (*ICD-9*) and *International Statistical Classification of Diseases, Tenth Revision* (*ICD-10*) diagnostic codes; and prescription data were extracted without time limits. An aggregate count of patients at each clinic meeting study eligibility, independent of nativity or language, was collected to determine the proportion study participants represented among the total population at each site. Descriptive analysis was completed in R (version 2025.05.01+513, 2025 Posit Software, PBC)

**Table 1.** Study Period Start and End Dates and Eligible Population by Site^a^, Tuberculosis Epidemiologic Studies Consortium (TBESC-III)

| Site<sup>b</sup> | Start date | End date | Age eligibility | Visit definition<br>(Any patient who has this type of visit at a clinic associated with the site is included) | Other eligibility criteria |
| --- | --- | --- | --- | --- | --- |
| Site A | 10/1/2020 | 9/30/2022 | At least 2 years of age at a visit that occurred during the study period. | In-person or telehealth in a primary care clinic. | NA |
| Site B | 10/1/2021 | 9/30/2022 | At least 18 years of age at a visit that occurred during the study period. | In-person office visit, video visit, or scheduled telephone visit with a physician (might be a resident or osteopath) in the Adult & Family Medicine Department. | At least 11 months of continuous membership in the health care network during the study period. |
| Site C | 10/1/2021 | 9/30/2022 | At least 18 years of age at a visit that occurred during the study period. | Office visit (in-person or telehealth) that is classified as a primary care or pulmonology visit. | NA |
| Site D | 12/1/2020 | 8/31/2022 | At least 18 years of age at a visit that occurred during the study period. | Preventative care, annual visit, or follow-up visit, not including patients who presented solely for urgent care, emergency visits, or prenatal care. | NA |

Abbreviation: NA, not applicable.
<sup>a</sup>Participating sites serve at least 10,000 non-US-born individuals annually from countries with high TB disease incidence rates (defined as $\geq 10$ cases per 100,000 persons among expatriates living in the US).
<sup>b</sup>For all sites, patients needed to be non-US-born or, if country of birth is missing, have a primary language other than English.

### Defining the LTBI Care Cascade Steps

The site percentage for each care cascade step was the count of participants who met the cascade step definition divided by the total number of participants who met the definition for the preceding step at that site. To account for a wide range of clinic population sizes and ensure percentages were not driven by a single site, the percentage of individuals completing each cascade step was reported as an average of site-specific percentages. Patients were required to complete all steps successively to be counted. Figure 1 summarizes the care cascade steps and their definitions. Time allowed for completing each step was based on site clinical practices; there was a consensus to allow for maximum time when there were differences in clinical practices among sites.

**Figure 1.**
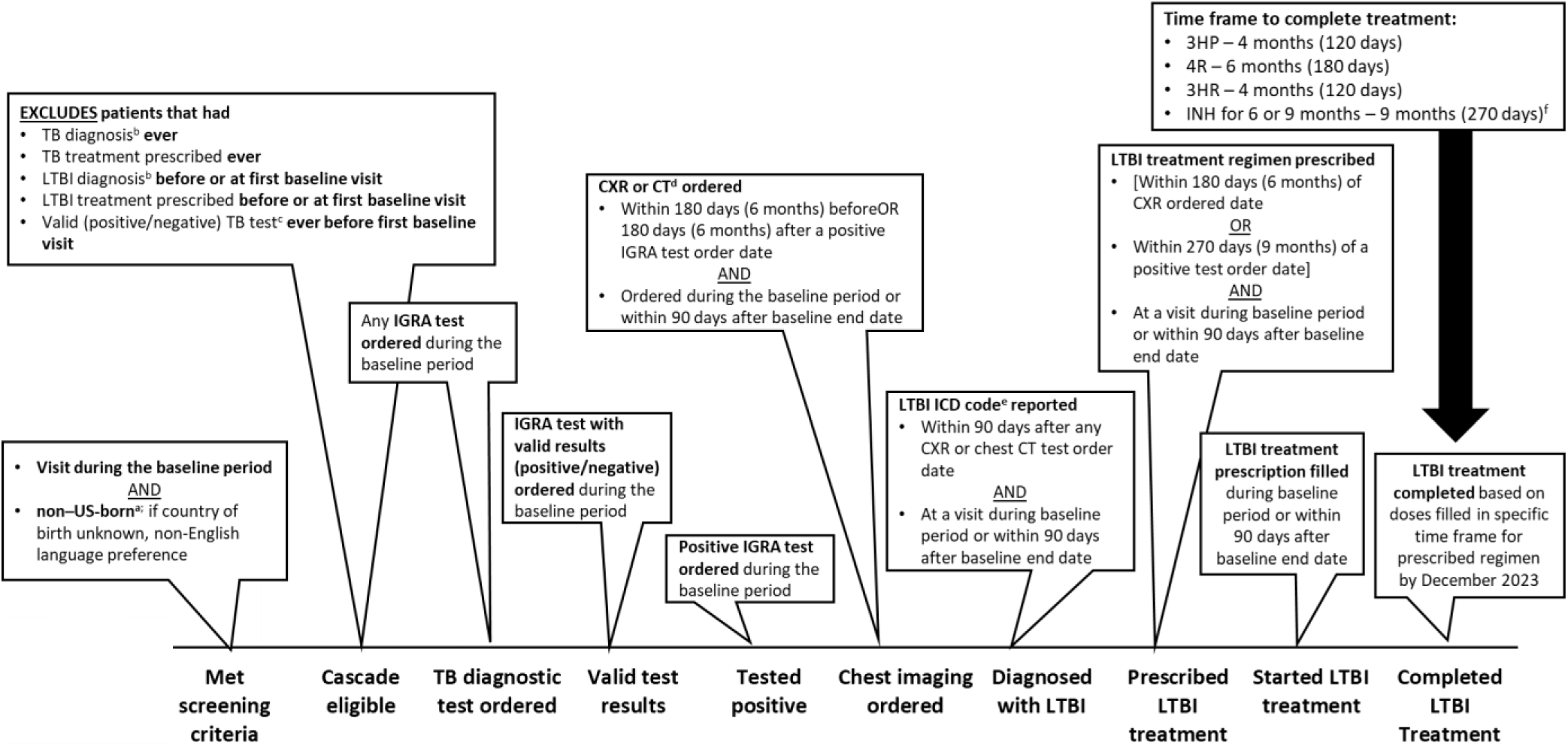
Latent Tuberculosis Infection Care Cascade Definitions, Tuberculosis Epidemiologic Studies Consortium (TBESC-III) ^a^Individuals born in Puerto Rico, US Virgin Islands, Guam, and the Northern Mariana Islands were categorized as US-born. ^b^Based on electronic health record data variables and *ICD-9* and *ICD-10* codes (eTable 1 in **Supplement 1**). ^c^Tuberculin skin test, IGRA, acid-fast bacilli culture or smear, or nucleic acid amplification test. A valid test result was defined as a positive or negative test result. ^d^CT data was submitted by 2 sites. ^e^ICD-9: 795.52 and ICD-10: R76.12 and Z22.7. ^f^If an individual completes 6 months of INH within 9 months, this amount of treatment would still be considered sufficient for treatment completion, even if they were prescribed 9 months of INH. 3HP indicates once-weekly isoniazid and rifapentine for 12 weeks; 3HR, 3 months of isoniazid and rifampin or rifabutin; 4R, 4 months of rifampin or rifabutin; CT, computed tomography; CXR, chest x-ray; *ICD-9*, *International Classification of Diseases*, *Ninth Revision*; *ICD-10*, *International Classification of Diseases, Tenth Revision;* IGRA, interferon-gamma release assay; INH, isoniazid; LTBI, latent tuberculosis infection; and TB, tuberculosis.

The care cascade begins with participants who met TB screening criteria, defined as reported non-US birth, or if country of birth was unknown or not collected, a primary language other than English. Demographic characteristics, specific risk factors for TB infection or disease, and insurance status at first visit during the study period were summarized for the population that met screening criteria; proportions for these variables were not averaged across sites but were stratified by site.

The next step, cascade eligibility, identified persons for whom screening was indicated based on having no evidence of prior testing or treatment. Persons with a TB or LTBI diagnosis or treatment prescribed before or at their first visit during the study period, or TB diagnosis or treatment prescribed during the study period, were ineligible. TB and LTBI diagnosis were defined based on submitted EHR visit variables and diagnostic codes (eTable 1 in **Supplement 1**). TB and LTBI treatment were based on submitted EHR visit variables and regimens prescribed using prescription data (eTable 2 in **Supplement 1**). We excluded individuals who had a valid diagnostic test for TB infection or disease before their first visit during the study period. A valid test result was defined as a positive or negative test result using a tuberculin skin test (TST), interferon-gamma release assay (IGRA), acid-fast bacilli culture or smear, or *Mycobacterium tuberculosis* complex nucleic acid amplification test.

Next, we identified persons with any IGRA test ordered during the study period, including TSPOT.*TB*, QuantiFERON-TB Gold, QuantiFERON-TB Gold In-Tube, or QuantiFERON-TB Gold Plus. We then determined those with a valid test result (positive or negative) and those with a positive test result. Next, we identified persons who had either a chest x-ray (CXR) or computed tomography (CT) of the chest ordered within 180 days before or after the date that a positive IGRA was ordered. This step was followed by an LTBI diagnosis (an LTBI *ICD-9* code [795.52] or *ICD-10* code [R76.12 and Z22.7]) reported within 90 days after the date any chest imaging was ordered. The CXR or CT had to be ordered and the diagnostic code reported during the study period or within 90 days after the study period end date for that site. Due to concerns that the lack of standardization across clinics in using LTBI diagnostic codes could underestimate the number of diagnoses, we also characterized a care cascade that removed the LTBI diagnosis step.

For the treatment steps, we identified the following LTBI treatment regimens using prescription drug records: isoniazid and rifapentine for 3 months (3HP), rifampin/rifabutin for 4 months (4R), isoniazid and rifampin/rifabutin for 3 months (3HR), and isoniazid for 6 months (6H) or 9 months (9H); we were unable to distinguish between 6H and 9H regimens. An LTBI treatment regimen had to be prescribed within 180 days of a chest imaging order date or 270 days of a positive diagnostic test order date; it had to be prescribed at a visit during the study period or within 90 days after the study period end date. To determine persons who started the treatment regimen, we assessed whether prescribed medications for a regimen were filled. Treatment completion was determined based on whether a minimum number of doses were filled within a specific time frame based on the prescribed regimen. Persons had until 14 months after the end of the study period to complete treatment. We stratified treatment prescribed, started, and completed by LTBI treatment regimen; multiple regimens could be prescribed and started for each individual. A detailed description of treatment prescription, initiation, and completion methods is available (eMethods 2 in **Supplement 1**).

### Ethical Considerations

This study was reviewed by and conducted under the authority of the CDC and was determined to not be human subjects research as the primary intent was routine disease surveillance. The study was conducted consistent with applicable federal law and CDC policy.^a^

## Results

There were 548,923 persons that met screening criteria and were included in the study (i.e. participants). This was an average of 47.9% of the total clinic population of 3,522,077 patients across sites. Among those who met screening criteria, the majority (52%; n=285,754) were aged 40 to 64 years; 58% (n=320,244) were female, and 37% (n=203,209) reported Hispanic ethnicity (Table 2). Among 326,993 persons who reported non-Hispanic ethnicity, the majority (82%; n=267,925) identified as Asian. Mexico was the most frequently reported country of birth among those who met screening criteria (20%; n=112,524), followed by China (15%; n=81,968) and the Philippines (12%; n=65,906). Among those who met screening criteria, the top 3 reported primary languages were English (49%; n=268,371), Spanish or Castilian (25%; n=138,721), and Chinese (15%; n=83,235).

**Table 2.**
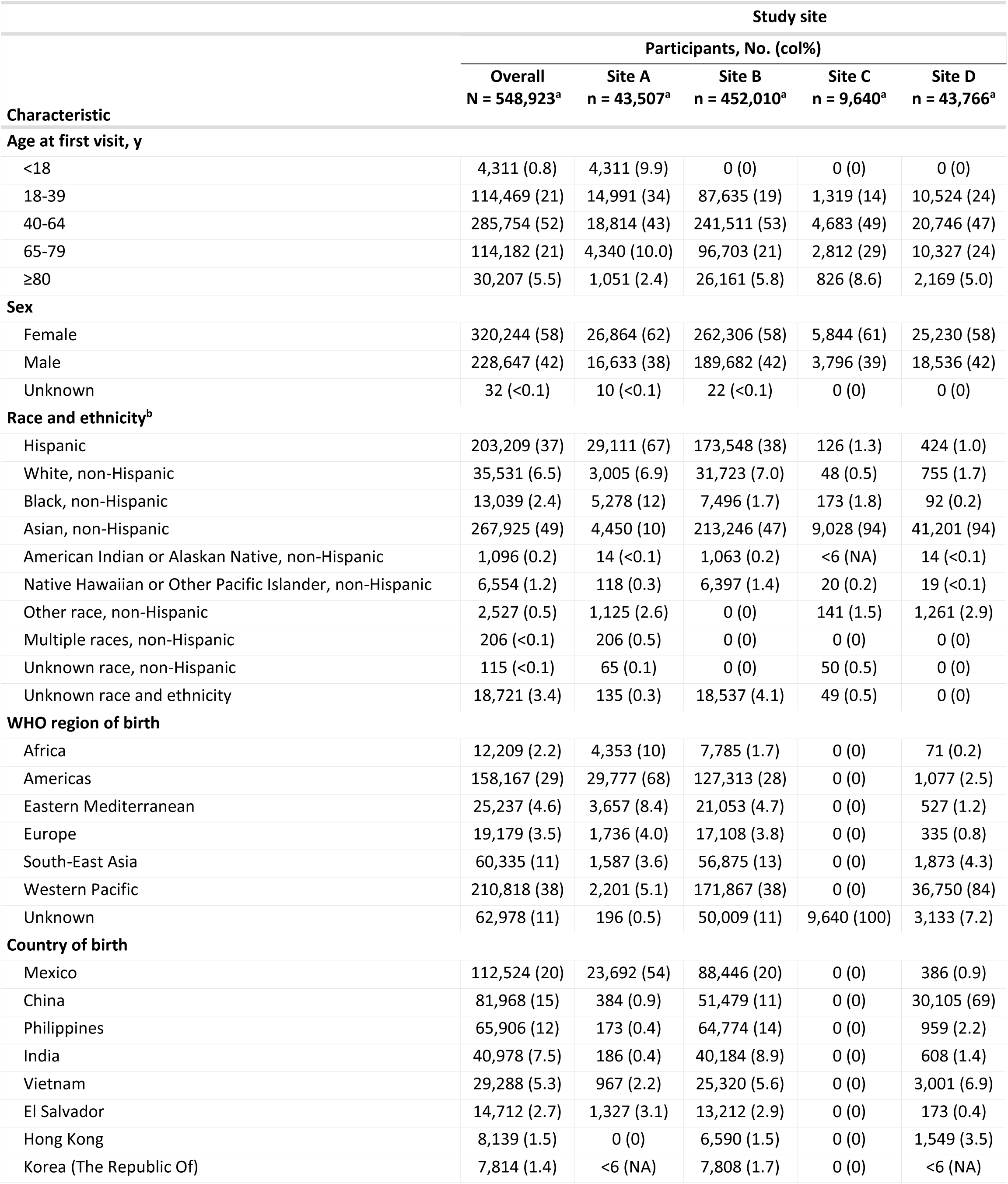
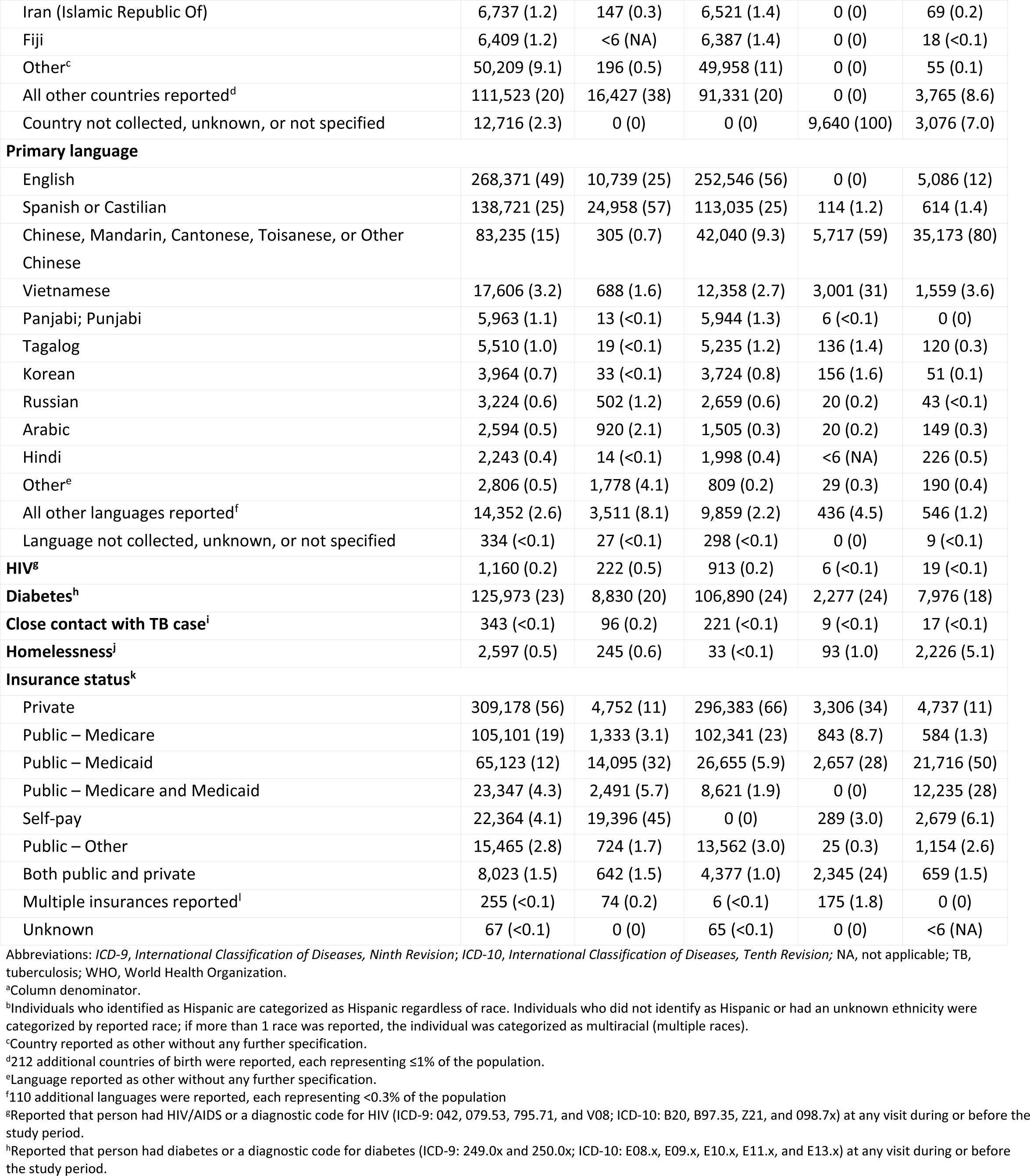
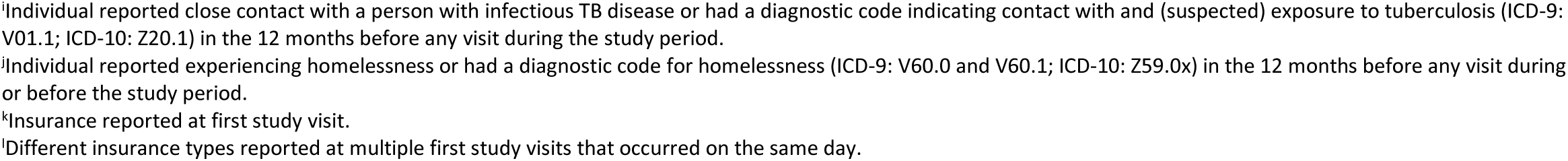
Characteristics of Participants Meeting Tuberculosis Screening Criteria by Study Site.

Among those who met screening criteria, an average of 68.8% (n=379,176) met cascade eligibility across sites (Figure 2). Among the 169,747 who were ineligible, the majority (90.7%; n=153,898) were removed due to having at least 1 TB diagnostic test with a positive or negative result before their first study period visit (eFigure 3 in **Supplement 1**). Regardless of testing history, 1.0% (n=1,653) of ineligible persons had evidence of TB diagnosis or treatment in their EHR, and 36.6% (n=62,133) had evidence of LTBI diagnosis or treatment before their first visit; 2.4% (n=4,140) had evidence of both TB disease and LTBI diagnosis or treatment.

**Figure 2.**
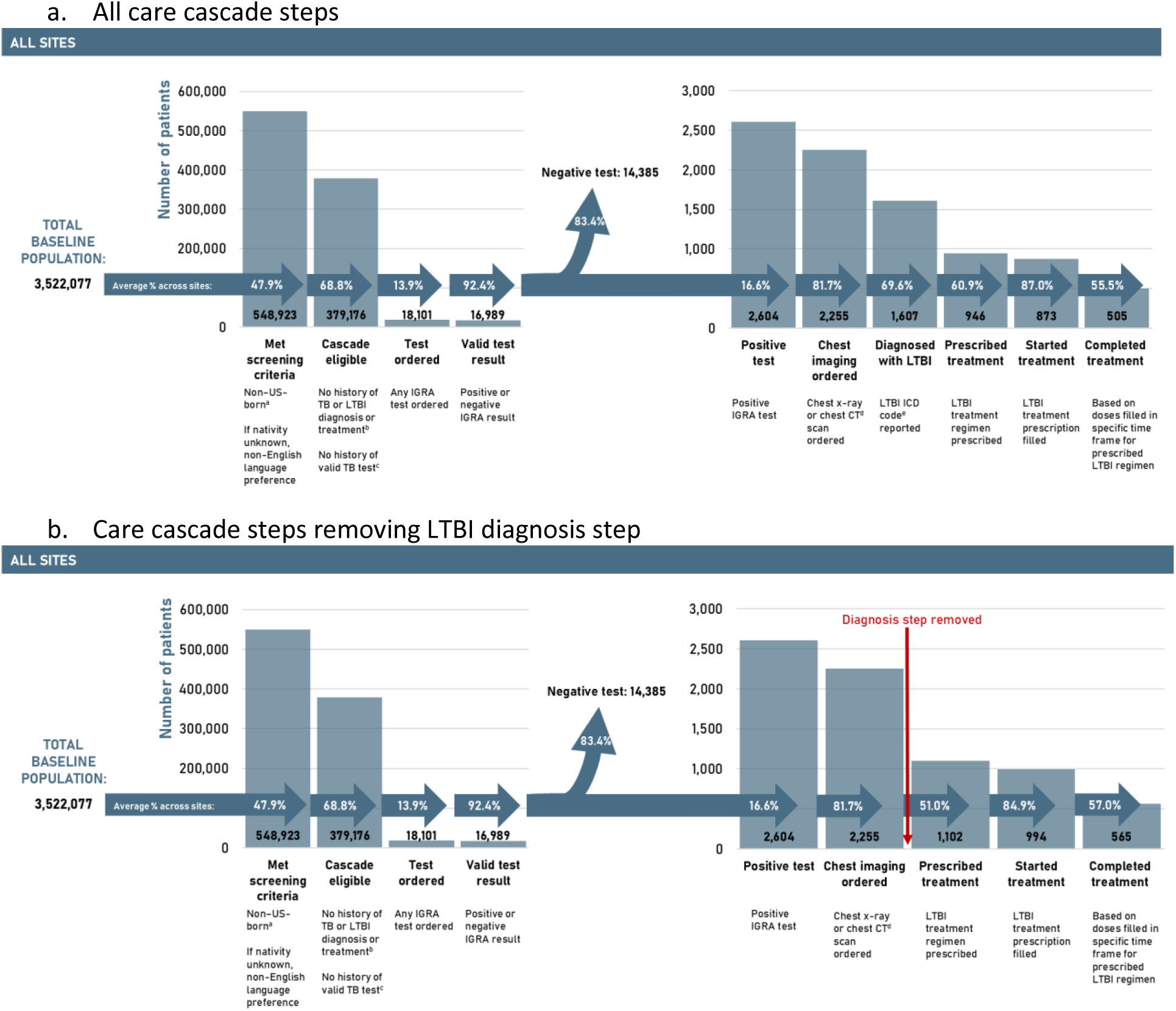
Latent Tuberculosis Infection Care Cascades in the Tuberculosis Epidemiologic Studies Consortium (TBESC-III) ^a^Individuals born in Puerto Rico, US Virgin Islands, Guam, and the Northern Mariana Islands were categorized as US-born. ^b^Based on electronic health record data and *ICD-9* and *ICD-10* codes. ^c^Tuberculin skin test, IGRA, acid-fast bacilli culture or smear, or nucleic acid amplification test. ^d^CT data was only submitted by 1 site. ^e^ICD-9: 795.52; ICD-10: R76.12 and Z22.7. CT indicates computed tomography; *ICD-9*, *International Classification of Diseases, Ninth Revision*; *ICD-10, International Classification of Diseases, Tenth Revision*; IGRA, interferon-gamma release assay; LTBI, latent tuberculosis infection; and TB, tuberculosis.

**Figure 3.**
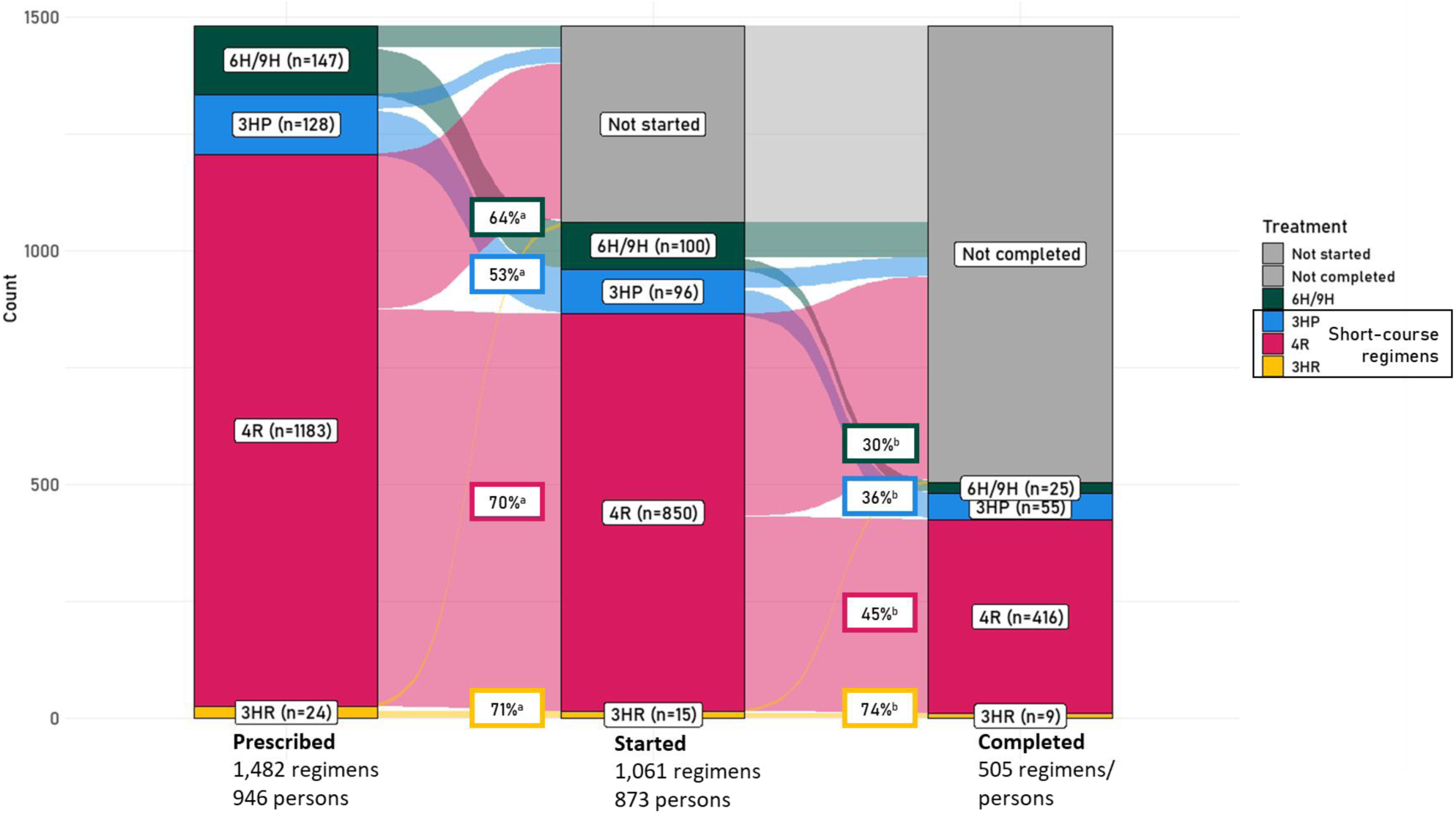
Latent Tuberculosis Infection Treatment Regimens Prescribed, Started, And Completed. ^a^Average percentage of regimens across sites prescribed that were started. ^b^Average percentage of regimens across sites started that were completed. 3HP indicates once-weekly isoniazid and rifapentine for 12 weeks; 3HR, 3 months of isoniazid and rifampin or rifabutin; 4R, 4 months of rifampin or rifabutin; 6H, isoniazid for 6 months; 9H, isoniazid for 9 months.

Among the cascade-eligible population, an average of 13.9% (n=18,101) had an IGRA ordered; an additional 0.36% (n=2,109), on average, were tested using TST. Among those tested using IGRA, an average of 92.4% (n=16,989) had a valid test result (i.e., positive or negative); an average of 16.6% (n=2,604) had a positive IGRA result, among those, an average of 81.7% (n=2,255) had chest imaging ordered. An average of 69.6% (n=1,607) were subsequently diagnosed with LTBI.

Among those diagnosed with LTBI, an average of 60.9% (n=946) were prescribed LTBI treatment. Overall, 1,482 regimens were prescribed among the 946 persons prescribed treatment; on average, 87.8% were rifamycin-based short-course regimens (Figure 3). Among those prescribed treatment, an average of 87.0% (n=873) started treatment; on average across sites, 69.5% of 4R, 52.9% of 3HP, 71.3% of 3HR, and 63.8% of 6H or 9H regimens prescribed were started. Among persons who started treatment, the average completion rate was 55.5% (n=505). On average, across sites, 45.3% of 4R, 35.6% of 3HP, 73.6% of 3HR, and 45.3% of 6H or 9H regimens started were completed.

In an alternate cascade removing the LTBI diagnostic step (Figure 2), among those with chest imaging, the average proportions were as follows: 51.0% (n=1,102) were prescribed treatment, 84.9% (n=994) started treatment, and 57.0% (n=565) completed treatment. Site-specific care cascades had notable differences across all care cascade steps (Figure 4).

**Figure 4.**
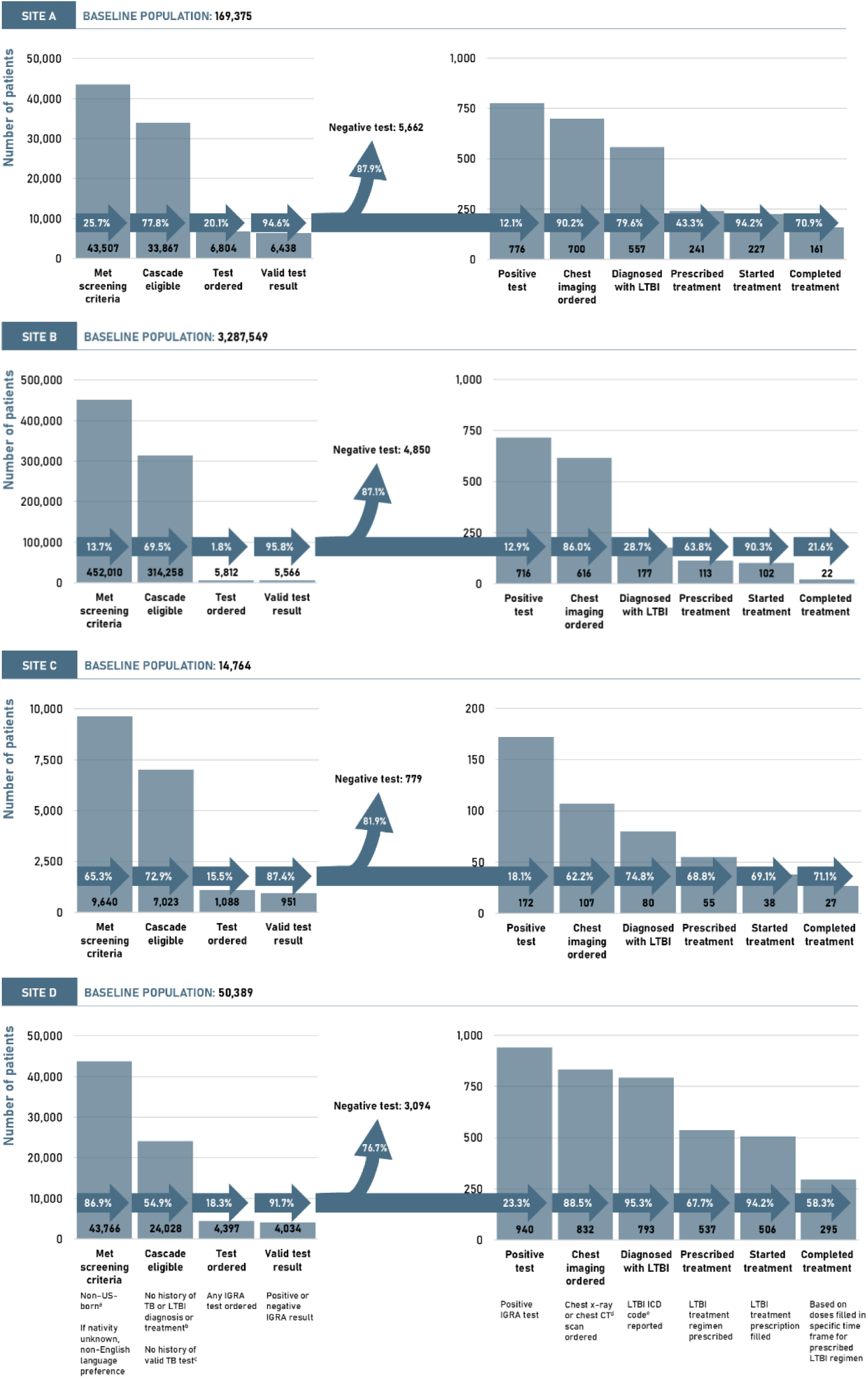
Latent Tuberculosis Infection Care Cascade by Tuberculosis Epidemiologic Studies Consortium (TBESC-III) Site. ^a^Individuals born in Puerto Rico, US Virgin Islands, Guam, and the Northern Mariana Islands were categorized as US-born. ^b^Based on electronic health record data and *ICD-9* and *ICD-10* codes. ^c^Tuberculin skin test, IGRA, acid-fast bacilli culture or smear, or nucleic acid amplification test. ^d^CT data was only submitted by 1 site. ^e^ICD-9: 795.52; ICD-10: R76.12 and Z22.7. CT indicates computed tomography; *ICD-9*, *International Classification of Diseases, Ninth Revision*; *ICD-10, International Classification of Diseases and Related Health Problems, Tenth Revision*; IGRA, interferon-gamma release assay; LTBI, latent TB infection; and TB, tuberculosis.

## Discussion

Using a longitudinal EHR cohort from 4 diverse primary care networks, we characterized the LTBI care cascade, from identifying those who met screening recommendations through treatment completion. Previous studies have characterized portions of the LTBI care cascade or analyzed specific populations, a single clinic or hospital system, but none have assessed the LTBI care cascade through treatment completion across multiple primary care settings. Nor have longitudinal EHR data elements and clinical protocols been used to establish timeframes for achieving steps.^14,27,32,33^ This LTBI care cascade establishes rates of LTBI testing, diagnosis, and treatment prescription, initiation, and completion among individuals at increased risk for LTBI who seek care in primary care settings. This analysis suggests opportunities to expand testing and treatment after LTBI diagnosis to prevent TB disease. Although an average of 17% of participants had TB infection, an average of 86% of higher-risk individuals were not tested. An average of 39% of individuals diagnosed with LTBI were not prescribed treatment, and nearly half (average, 44%) did not finish treatment.

Primary care clinics are relevant settings for characterizing the LTBI care cascade and addressing areas where TB prevention can be expanded along the care cascade. Almost half the population seeking care at these primary care clinics were identified as being at higher risk for TB infection. Additionally, the average 16.6% IGRA test positivity observed is similar to other reported LTBI prevalence rates among non–US-born persons based on IGRA positivity.^34–38^

Previous studies have demonstrated opportunities to prevent TB disease through assessment of LTBI care cascade outcomes, including testing persons recommended for screening with recommended diagnostic tests, prescribing treatment for persons diagnosed, and ensuring treatment completion.^7,13–16,39^ Similarly, this cascade demonstrates the potential for improving screening among populations at higher risk and treatment outcomes among those diagnosed with LTBI. The proportion tested was lower than reported in other primary care^13,14,39^ and nonprimary care settings,^15^ among populations at higher risk for infection. However, using IGRA predominantly for TB testing contrasts with previous studies in primary care settings, where TST was predominantly used.^14,26,40,41^

We found a higher proportion of treatment prescribed and subsequently started compared with other studies,^15^ including those in other primary care settings.^14,16,39^ Although the proportion that completed treatment was higher than in similar settings,^14–16,42^ it was lower than among individuals who received care at public health clinics^21,26,43^ or where directly observed therapy was performed.^44^ The rates of treatment prescription, initiation, and completion varied across sites and treatment regimens.

Most treatment regimens prescribed were rifamycin-based regimens, indicating that TBESC-III sites have already implemented CDC’s recommendation for short-course rifamycin-based regimens, in contrast with other studies in primary care settings that show many practitioners might still widely use 6H/9H.^14,16,44^ As seen in other studies, we observed that short-course regimens had higher completion rates than 6H/9H.^7,15,44,45^ The high proportion of short-course regimens prescribed might have contributed to the higher rate of LTBI treatment completion in these primary care settings.

We developed standardized methods for defining every step in the LTBI care cascade using EHR data, including establishing time allowed for achieving steps based on sites’ clinical protocol, and detailed methodology for determining treatment prescription, initiation, and completion. The EHR data elements and definitions for each step of this care cascade are the first to incorporate the longitudinal progression of care (e.g., diagnosis after chest imaging, treatment prescription after diagnosis) in totality, from screening to treatment completion.

However, challenges in defining care cascade steps persisted. The number of individuals who met screening criteria was based on imperfect factors. Preferred language was a proxy for country of birth for all individuals at the smallest study site, and EHR data did not include travel or prior residence in countries outside the US. We were also unable to assess other risk factors for TB, including immunosuppression, close contact with a TB case, or history of homelessness or incarceration, when determining if an individual met screening criteria. In addition, CXR results were unavailable, so diagnostic codes were used as a proxy for LTBI diagnosis after TB disease was ruled out. However, LTBI diagnostic code use varies widely across clinics and physicians.^46^ Previous literature has shown that diagnostic codes alone have low sensitivity when trying to identify TB cases in EHR records.^29,47^ Similarly, TBESC-III clinics indicated LTBI diagnostic codes used to define the diagnostic step of the care cascade might not fully capture all persons diagnosed with LTBI. Our alternate care cascade that removed the LTBI diagnostic code requirement indicated an additional 6.9% (n=156) of persons who received chest imaging were prescribed treatment despite not having an LTBI diagnostic code present in the patient’s EHR. Thus, requiring an LTBI diagnostic code might underestimate LTBI diagnosis and treatment.

These sites are not representative of all primary care clinics in the US. This analysis was of a self-selected group of clinics dedicated to improving TB, evidenced by their desire to join TBESC-III. Thus, the uptake of TB testing and LTBI treatment recommendations in other primary care settings could be lower elsewhere. Additionally, each clinic had different clinical practices surrounding steps of the care cascade, specifically related to the time between achieving each step. Thus, although the consortium came to a consensus for time allowed between events, all individuals moving through the care cascade may not have been identified.

Several limitations were identified related to determining treatment initiation and completion based on pharmacy data. We were unable to determine from EHR data if individuals were offered treatment and declined it. Furthermore, prescription variables, such as dosing, frequency, quantity, and refills, were not standardized across sites and were sometimes incomplete, requiring an assumption that common dosing and frequency were used based on CDC guidelines.^7^ Individuals might also have filled prescriptions at outside pharmacies, causing an underestimation of treatment initiation and completion. Lastly, we were unable to confirm whether individuals actually ingested the medication.

Finally, EHR data available might not represent the entirety of an individual’s medical history. Previous testing, diagnosis, or treatment for LTBI or TB disease might not have been available, and individuals might have been inappropriately deemed eligible for testing; this could have downstream effects on the care cascade.

In summary, we established a standardized methodology for estimating a complete LTBI care cascade using longitudinal EHR data, including standardized time intervals between steps and a new approach to assess LTBI treatment prescription, initiation, and completion using pharmacy data. We used these methods to assess outcomes in primary care settings where a large proportion of the population met TB screening criteria. We identified opportunities for expanding TB prevention efforts in TB testing, and in prescribing and completing LTBI treatment. Using this information, health care providers can design and implement interventions to prevent TB-associated morbidity and advance TB elimination in the US. This approach can be applied in other primary care settings to identify opportunities for improving TB care, as well as to design and evaluate the impact of specific interventions.

## Data Availability

All data produced in the present study will be made publicly available after the conclusion of the contract.

## Acknowledgments

We would like to acknowledge the contributions of Kumar Batra, Danique Gigger, Juan (Antonio) Hernandez, Mehabuba Rahman, Julia Raykin, Noah Schwartz, Sammi Smith, Thara Venkatappa, Jonathan Wortham, and Angus Wu to this manuscript. Furthermore, we thank all study participants and site staff who are participating in TBESC-III.

## Footnotes

a See 45 C.F.R. part 46, 21 C.F.R. part 56; 42 U.S.C. §241(d); 5 U.S.C. §552a; 44 U.S.C. §3501 et seq.

## References

1. Filardo TD, Feng PJ, Pratt RH, Price SF, Self JL. Tuberculosis - United States, 2021. MMWR Morb Mortal Wkly Rep. 2022;71(12):441–446. doi:10.15585/mmwr.mm7112a1

2. Williams PM, Pratt RH, Walker WL, Price SF, Stewart RJ, Feng PJI. Tuberculosis — United States, 2023. MMWR Morb Mortal Wkly Rep. 2024;73(12):265–270.

3. Shea KM, Kammerer JS, Winston CA, Navin TR, Horsburgh CR. Estimated rate of reactivation of latent tuberculosis infection in the United States, overall and by population subgroup. Am J Epidemiol. Published online 2014. doi:10.1093/aje/kwt246

4. Yuen CM, Kammerer JS, Marks K, Navin TR, France AM. Recent Transmission of Tuberculosis — United States, 2011–2014. PLoS One. 2016;11(4):e0153728. doi:10.1371/journal.pone.0153728

5. Menzies NA, Cohen T, Hill AN, et al. Prospects for tuberculosis elimination in the United States: Results of a transmission dynamic model. Am J Epidemiol. Published online 2018. doi:10.1093/aje/kwy094

6. Lobue P, Menzies D. Treatment of latent tuberculosis infection: An update. Respirology. Published online 2010. doi:10.1111/j.1440-1843.2010.01751.x

7. Sterling TR, Njie G, Zenner D, et al. Guidelines for the Treatment of Latent Tuberculosis Infection: Recommendations from the National Tuberculosis Controllers Association and CDC, 2020. MMWR Morb Mortal Wkly Rep. Published online 2020. doi:10.1111/ajt.15841

8. International Union Against Tuberculosis Committee on Prophylaxis. Efficacy of various durations of isoniazid preventive therapy for tuberculosis: five years of follow-up in the IUAT trial. International Union Against Tuberculosis Committee on Prophylaxis. Bull World Health Organ. 1982;60(4):555–564.

9. Collins JM, Stout JE, Ayers T, et al. Prevalence of Latent Tuberculosis Infection Among Non-US-Born Persons by Country of Birth-United States, 2012-2017. Clin Infect Dis an Off Publ Infect Dis Soc Am. 2021;73(9):e3468–e3475. doi:10.1093/cid/ciaa1662

10. Lewinsohn DM, Leonard MK, Lobue PA, et al. Official American Thoracic Society/Infectious Diseases Society of America/Centers for Disease Control and Prevention Clinical Practice Guidelines: Diagnosis of Tuberculosis in Adults and Children. Clin Infect Dis. Published online 2017. doi:10.1093/cid/ciw694

11. US Preventive Services Task Force. Screening for Latent Tuberculosis Infection in Adults: US Preventive Services Task Force Recommendation Statement. JAMA. 2016;316(9):962–969. doi:10.1001/jama.2016.11046

12. Centers for Disease Control and Prevention (CDC). Latent Tuberculosis Infection: A Guide for Primary Health Care Providers.; 2020. https://www.cdc.gov/tb/publications/ltbi/pdf/LTBIbooklet508.pdf

13. Tang AS, Mochizuki T, Dong Z, Flood J, Katrak SS. Can Primary Care Drive Tuberculosis Elimination? Increasing Latent Tuberculosis Infection Testing and Treatment Initiation at a Community Health Center with a Large Non-U.S.-born Population. J Immigr Minor Heal. 2023;25(4):803–815. doi:10.1007/s10903-022-01438-1

14. Vonnahme LA, Raykin J, Jones M, et al. Using Electronic Health Record Data to Measure the Latent Tuberculosis Infection Care Cascade in Safety-Net Primary Care Clinics. AJPM Focus. 2023;2(4):100148. 10.1016/j.focus.2023.100148

15. Alsdurf H, Hill PC, Matteelli A, Getahun H, Menzies D. The cascade of care in diagnosis and treatment of latent tuberculosis infection: a systematic review and meta-analysis. Lancet Infect Dis. Published online 2016. doi:10.1016/S1473-3099(16)30216-X

16. Bruxvoort KJ, Skarbinski J, Fischer H, et al. Latent Tuberculosis Infection Treatment Practices in Two Large Integrated Health Systems in California, 2009–2018. Open Forum Infect Dis. 2023;10(5):ofad219. doi:10.1093/ofid/ofad219

17. Tasillo A, Salomon JA, Trikalinos TA, Horsburgh CRJ, Marks SM, Linas BP. Cost-effectiveness of Testing and Treatment for Latent Tuberculosis Infection in Residents Born Outside the United States With and Without Medical Comorbidities in a Simulation Model. JAMA Intern Med. 2017;177(12):1755–1764. doi:10.1001/jamainternmed.2017.3941

18. Chatterji M, Donald M, van Driel M, Marks G, Liaw S, Sharman L. Models of care for tuberculosis infection screening and treatment in primary care: A scoping review. Aust J Gen Pract. 2024;53:756–763. https://www1.racgp.org.au/ajgp/2024/october/models-of-care-for-tuberculosis-infection-screenin

19. Carter KL, Gabrellas AD, Shah S, Garland JM. Improved latent tuberculosis therapy completion rates in refugee patients through use of a clinical pharmacist. Int J Tuberc lung Dis Off J Int Union against Tuberc Lung Dis. 2017;21(4):432–437. doi:10.5588/ijtld.16.0575

20. D’Lugoff MI, Jones W, Kub J, et al. Tuberculosis screening in an at-risk immigrant Hispanic population in Baltimore city: an academic health center/local health department partnership. J Cult Divers. 2002;9(3):79–85.

21. Prater C, Holzman S, Shah M. Programmatic Effectiveness of Latent Tuberculosis Care Cascade in a Community Health Center. J Immigr Minor Heal. 2021;23(3):566–573. doi:10.1007/s10903-020-01047-w

22. Steele AW, Eisert S, Davidson A, et al. Using computerized clinical decision support for latent tuberculosis infection screening. Am J Prev Med. 2005;28(3):281–284. doi:10.1016/j.amepre.2004.12.012

23. Ehman M, Flood J, Barry PM. Tuberculosis Treatment Managed by Providers outside the Public Health Department: Lessons for the Affordable Care Act. PLoS One. 2014;9(10):e110645-. 10.1371/journal.pone.0110645

24. Arsenault C, Roder-DeWan S, Kruk ME. Measuring and improving the quality of tuberculosis care: A framework and implications from the Lancet Global Health Commission. J Clin Tuberc Other Mycobact Dis. 2019;16:100112. 10.1016/j.jctube.2019.100112

25. Kruk ME, Gage AD, Arsenault C, et al. High-quality health systems in the Sustainable Development Goals era: time for a revolution. Lancet Glob Heal. 2018;6(11):e1196–e1252. 10.1016/S2214-109X(18)30386-3

26. Holzman SB, Perry A, Saleeb P, et al. Evaluation of the Latent Tuberculosis Care Cascade Among Public Health Clinics in the United States. Clin Infect Dis. Published online April 1, 2022:ciac248. doi:10.1093/cid/ciac248

27. Jenks J, Garfein R, Zhu W, Hogarth M. Latent Tuberculosis Screening Using Electronic Health Record Data. Emerg Infect Dis J. 2020;26(9):2285. doi:10.3201/eid2609.191391

28. Vonnahme LA. Novel Tools to Measure Latent Tuberculosis Infection Among Populations at Higher Risk in the United States. Georgia State University; 2024. 10.57709/36275860

29. Birkhead GS, Klompas M, Shah NR. Uses of Electronic Health Records for Public Health Surveillance to Advance Public Health. Annu Rev Public Health. Published online 2015. doi:10.1146/annurev-publhealth-031914-122747

30. Centers for Disease Control and Prevention (CDC). Tuberculosis Epidemiologic Studies Consortium. Published 2024. Accessed June 12, 2024. https://www.cdc.gov/tb/research/tbesc.html

31. Tsang CA, Langer AJ, Steve Kammerer J, Navin TR. US tuberculosis rates among persons born outside the United States compared with rates in their countries of birth, 2012-2016. Emerg Infect Dis. Published online 2020. doi:10.3201/eid2603.190974

32. Stockbridge EL, Loethen AD, Annan E, Miller TL. Interferon gamma release assay tests are associated with persistence and completion of latent tuberculosis infection treatment in the United States: Evidence from commercial insurance data. PLoS One. 2020;15(12):e0243102. doi:10.1371/journal.pone.0243102

33. Stockbridge EL, Miller TL, Carlson EK, Ho C. Tuberculosis Prevention in the Private Sector: Using Claims-Based Methods to Identify and Evaluate Latent Tuberculosis Infection Treatment With Isoniazid Among the Commercially Insured. J Public Health Manag Pract. 2018;24(4):E25–E33. doi:10.1097/PHH.0000000000000628

34. Yelk Woodruff R, Hill A, Marks S, Navin T, Miramontes R. Estimated Latent Tuberculosis Infection Prevalence and Tuberculosis Reactivation Rates Among Non-U.S.-Born Residents in the United States, from the 2011-2012 National Health and Nutrition Examination Survey. J Immigr Minor Heal. 2021;23(4):806–812. doi:10.1007/s10903-020-01065-8

35. Miramontes R, Hill AN, Yelk Woodruff RS, et al. Tuberculosis Infection in the United States: Prevalence Estimates from the National Health and Nutrition Examination Survey, 2011-2012. PLoS One. 2015;10(11):e0140881. doi:10.1371/journal.pone.0140881

36. Mancuso JD, Diffenderfer JM, Ghassemieh BJ, Horne DJ, Kao TC. The prevalence of latent tuberculosis infection in the United States. Am J Respir Crit Care Med. Published online 2016. doi:10.1164/rccm.201508-1683OC

37. Vonnahme LA, Haddad MB, Navin TR. Factoring prior treatment into tuberculosis infection prevalence estimates, United States, 2011-2012. Emerg Infect Dis. Published online 2019. doi:10.3201/eid2510.190439

38. Mirzazadeh A, Kahn JG, Haddad MB, et al. State-level prevalence estimates of latent tuberculosis infection in the United States by medical risk factors, demographic characteristics and nativity. PLoS One. 2021;16(4):e0249012. doi:10.1371/journal.pone.0249012

39. Shapiro AE, Gupta A, Lan K, Kim HN. Latent Tuberculosis Screening Cascade for Non–US-Born Persons in a Large Health System. Open Forum Infect Dis. 2023;10(7):ofad303. doi:10.1093/ofid/ofad303

40. Stockbridge EL, Miller TL, Carlson EK, Ho C. Private sector tuberculosis prevention in the US: Characteristics associated with interferon-gamma release assay or tuberculin skin testing. PLoS One. 2018;13(3):e0193432. 10.1371/journal.pone.0193432

41. Owusu-Edusei K, Stockbridge EL, Winston CA, Kolasa M, Miramontes R. Tuberculin skin test and interferon-gamma release assay use among privately insured persons in the United States. Int J Tuberc lung Dis Off J Int Union against Tuberc Lung Dis. 2017;21(6):684–689. doi:10.5588/ijtld.16.0617

42. Horsburgh CR, Goldberg S, Bethel J, et al. Latent TB infection treatment acceptance and completion in the United States and Canada. Chest. Published online 2010. doi:10.1378/chest.09-0394

43. Nuzzo JB, Golub JE, Chaulk P, Shah M. Postarrival Tuberculosis Screening of High-Risk Immigrants at a Local Health Department. Am J Public Health. 2015;105(7):1432–1438. doi:10.2105/AJPH.2014.302287

44. Lines G, Hunter P, Bleything S. Improving Treatment Completion Rates for Latent Tuberculosis Infection: A Review of Two Treatment Regimens at a Community Health Center. J Health Care Poor Underserved. 2015;26(4):1428–1439. doi:10.1353/hpu.2015.0126

45. Njie GJ, Morris SB, Woodruff RY, Moro RN, Vernon AA, Borisov AS. Isoniazid-Rifapentine for Latent Tuberculosis Infection: A Systematic Review and Meta-analysis. Am J Prev Med. 2018;55(2):244–252. 10.1016/j.amepre.2018.04.030

46. Iqbal SA, Isenhour CJ, Mazurek G, Truman BI. Diagnostic code agreement for electronic health records and claims data for tuberculosis. Int J Tuberc lung Dis Off J Int Union against Tuberc Lung Dis. 2020;24(7):706–711. doi:10.5588/ijtld.19.0792

47. Calderwood MS, Platt R, Hou X, et al. Real-time surveillance for tuberculosis using electronic health record data from an ambulatory practice in eastern Massachusetts. Public Health Rep. 2010;125(6):843–850. doi:10.1177/003335491012500611

