## Supplemental Materials for "Measuring the Latent Tuberculosis Infection Care Cascade Using Electronic Health Record Data from Primary Care Clinics in the Tuberculosis Epidemiologic Studies Consortium-III"

**Supplement 1**

**eMethods 1. TBESC-III Study Design and Population**

The Tuberculosis Epidemiologic Studies Consortium (TBESC) is a CDC-funded collaboration with academic, public health, and primary care partners and is in its third iteration (TBESC-III).[1] The current cycle began in September 2021 and is scheduled to end in March 2026. The mission of TBESC-III is to assist tuberculosis (TB) elimination efforts in the US by focusing on latent tuberculosis infection (LTBI) among non–US-born populations at higher risk seeking care in primary care settings. The objective of the consortium is to use implementation science to identify primary care interventions that increase LTBI targeted testing and treatment and that are effective and efficient. Four TBESC-III sites identified as primary care settings serving non–US-born persons were selected to design and implement clinical and patient-centered interventions to improve performance measures across the LTBI care cascade. The consortium study design includes a baseline and intervention period; for the purposes of this analysis, the study period only includes the baseline period. The baseline period was required to be a minimum of 12 months immediately preceding the implementation of the intervention(s), with eligible individuals being those who sought care during this time. The baseline period was chosen by each site, with some sites choosing a longer baseline; each site also had varying age eligibility criteria and primary care visit definitions (Table 1 in main manuscript). The intervention period starts immediately afterwards and includes all eligible individuals who seek care during that time.

To monitor and evaluate intervention performance over time, sites collect retrospective and prospective electronic health record (EHR) data on the study cohort. Comprehensive EHR data on all individuals in the study cohort are submitted every 3 months until the end of the consortium to ensure data for the study cohort are up to date and to track intervention progress. Due to large clinic populations and cumulative data, 3 of 4 sites extracted patient-level EHR data only for the subset who met screening guidelines based on non-US birth or non-English language as their primary language if country of birth was missing, rather than the entire clinic population; aggregated information on the total clinic population was submitted separately by these sites. The dataset for the baseline period was finalized after data were extracted in December 2023, approximately 14 months after the baseline period concluded (depending on the site). This time frame allowed for persons who were screened at the end of the baseline period to progress through the entire LTBI care cascade, allowing for sufficient time to complete LTBI treatment, if relevant, and ensuring that their EHR was updated to reflect all clinical updates. This dataset was used for this analysis.

**eTable 1. Tuberculosis Disease and Latent Tuberculosis Infection Definitions for Determining Latent Tuberculosis Care Cascade Eligibility**

| **Condition** | **Definition for determining ineligibility for the LTBI care cascade** | ***ICD-9* diagnostic codes** | ***ICD-10* diagnostic codes** |
|---|---|---|---|
| Tuberculosis disease | ≥1 diagnostic code at any visit in the patient's electronic health record. | 010, 011, 012, 013, 014, 015, 016, 017, and 018 | A15.x, A17.x, A18.x, and A19.x |
| LBTI | ≥1 diagnostic code at any visit prior to or at the patient's first visit within the study period in their electronic health record. | 795.51 and 795.52 | R76.11, R76.12, and Z22.7 |

Abbreviations: *ICD-9*, *International Classification of Diseases, Ninth Revision*; *ICD-10, International Classification of Diseases, Tenth Revision*; LTBI, latent tuberculosis infection.

**eTable 2. Treatment Regimens for Tuberculosis Disease and Latent Tuberculosis Infection**

| TB regimens | LTBI regimens |
|---|---|
| INH + RIF + PZA + EMB | INH only |
| INH + PZA + RPT + Moxifloxacin | RIF only |
| INH + RIF + PZA | INH + RIF |
| INH + RIF + EMB | INH + RPT |
| INH + RIF + PZA + EMB + Levofloxacin | |
| INH + Rifampin + Rifabutin + PZA + EMB + Levofloxacin | |
| INH + RIF + PZA + EMB + Moxifloxacin | |
| RIF + PZA + EMB + Levofloxacin | |
| INH + RIF + PZA + EMB + Levofloxacin + Cycloserine | |
| Bedaquiline + Pretomanid + Linezolid | |

Abbreviations: EMB, ethambutol; INH, isoniazid; PZA, pyrazinamide; RIF, rifampin or rifabutin unless otherwise indicated; RPT, rifapentine.

**eMethods 2. Determining Treatment Prescription, Initiation, and Completion Using Electronic Health Record Data Elements in TBESC-III**

We applied a structured approach for analyzing prescription and regimen data from electronic health record (EHR) data to identify individuals who were prescribed and started tuberculosis (TB) and latent tuberculosis infection (LTBI) regimens, as well as to assess individuals' success in completing these regimens (eFigure 1). First, all medication prescriptions used individually or in combination to treat either TB or LTBI (eTable 2) were grouped into a prescription group if they were prescribed on the same day for that individual.

Since rifampin and rifabutin (hereafter both referred to as RIF) can be used to treat other conditions,[2] a previously published algorithm[3] was expanded upon to determine whether RIF prescriptions were for treatment of TB disease, LTBI, or another condition (eFigure 2). Each RIF prescription was assessed as being a part of a TB or LTBI regimen by evaluating other medications prescribed on the same day, the prescribed frequency of the RIF prescription, and the *International Classification of Diseases, Ninth Revision* (*ICD-9*) or *International Classification of Diseases, Tenth Revision* (*ICD-10*) diagnostic codes reported within the 60 days before the RIF prescription. RIF prescriptions were determined **not** to be prescribed as part of a TB or LTBI regimen under the following conditions:

1. RIF was prescribed in the absence of isoniazid or pyrazinamide on the same day *AND*
    a. At a frequency of greater than once per day *OR*
    b. Prescribed at a frequency once per day *AND* the individual had a diagnostic code in the 60 days before a condition not related to TB or LTBI determined to be treated using RIF (eTable 3) *WITHOUT* a TB or LTBI code in the preceding 60 days.

All RIF prescriptions meeting the above criteria were subsequently dropped. All other RIF prescriptions were maintained within their prescription groups, and the RIF prescription was deemed part of a TB or

LTBI regimen. All prescription groups were then assigned a TB or LTBI regimen based on the medications contained in the prescription group (eTable 4). Individuals with an assigned LTBI regimen were considered to have been prescribed LTBI treatment. Individuals with a TB regimen(s) were determined to be ineligible for the LTBI care cascade.

Once each prescription group was assigned a regimen, a treatment group was subsequently created based on consecutive pharmacy fills that aligned with the regimen's medications and recommended duration of the regimen. If a pharmacy refill was 62 days or less from a previous refill and the total duration of the regimen was less than or equal to the maximum time allotted for the specified regimen (eTable 5), then the refills were grouped together to create a treatment group. If a treatment group was created for a prescription group, the individual was considered to have started a regimen. Any treatment groups with a RIF prescription were then reassessed and confirmed to be a TB or LTBI regimen if at least 1 RIF prescription in the treatment group had been previously identified as prescribed for a TB or LTBI regimen or if the treatment group with the RIF prescription was determined to be ≥30 days duration.

Prescription frequency, pill dosage, and pill quantity from the EHR data were used to determine whether a regimen was completed. The pharmacy was required to be on the same EHR system as the site for these data to be available. Due to missingness and nonstandardization in values, prescription frequency and pill dosage were imputed if missing based on expected values for the assigned medication and regimen[4] (eTable 5). Pill dosage and pill quantity were used in a calculation to determine the number of doses per month and the monthly dosage (mg) of a prescription.

After calculating the prescription dosages for a medication, the dosages were summed across the treatment group to determine the total amount of medication taken by the individual. Treatment completion status (complete, incomplete, or in progress) was assigned based on the total dose count and total dosage of each medication taken for the regimen relative to the calculated minimum numbers of doses and total minimum dosage for completion of each medication in the regimen (eTable 5). The minimum number of doses and minimum total dosage to define treatment completion were derived by multiplying the total expected number of doses and total expected dosage for an adult[4] by 80%.[5–11] Each regimen was also evaluated against time constraints to determine if it met the minimum duration allowed and did not exceed the maximum regimen duration allowed[4].

Furthermore, Site D completed a manual chart review to evaluate regimen assignment and treatment completion. Data from the manual review were incorporated into the logic to compare the manual review with the treatment logic determinations for each individual to supplement missed regimens or completed statuses. At the study participant level, 97.1% concordance was observed between the applied structured approach for analyzing prescription and regimen data from the EHR and the chart review when determining the LTBI regimen assigned. Additionally, incorporating the manual chart review data improved treatment completion rates by 29.2%.

**eFigure 1. Framework for Determining Treatment Prescribed, Started, And Completed Using Electronic Health Record Data**

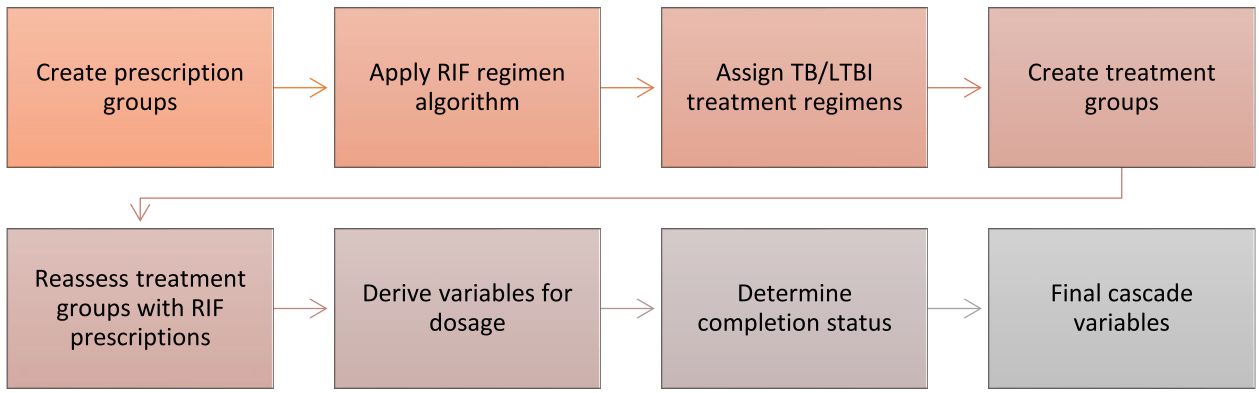


LTBI indicates latent tuberculosis infection; RIF, rifampin or rifabutin; and TB, tuberculosis.

### eFigure 2. Determining Whether Rifampin or Rifabutin Prescriptions are Part of Tuberculosis Disease or Latent Tuberculosis Infection Treatment Regimens

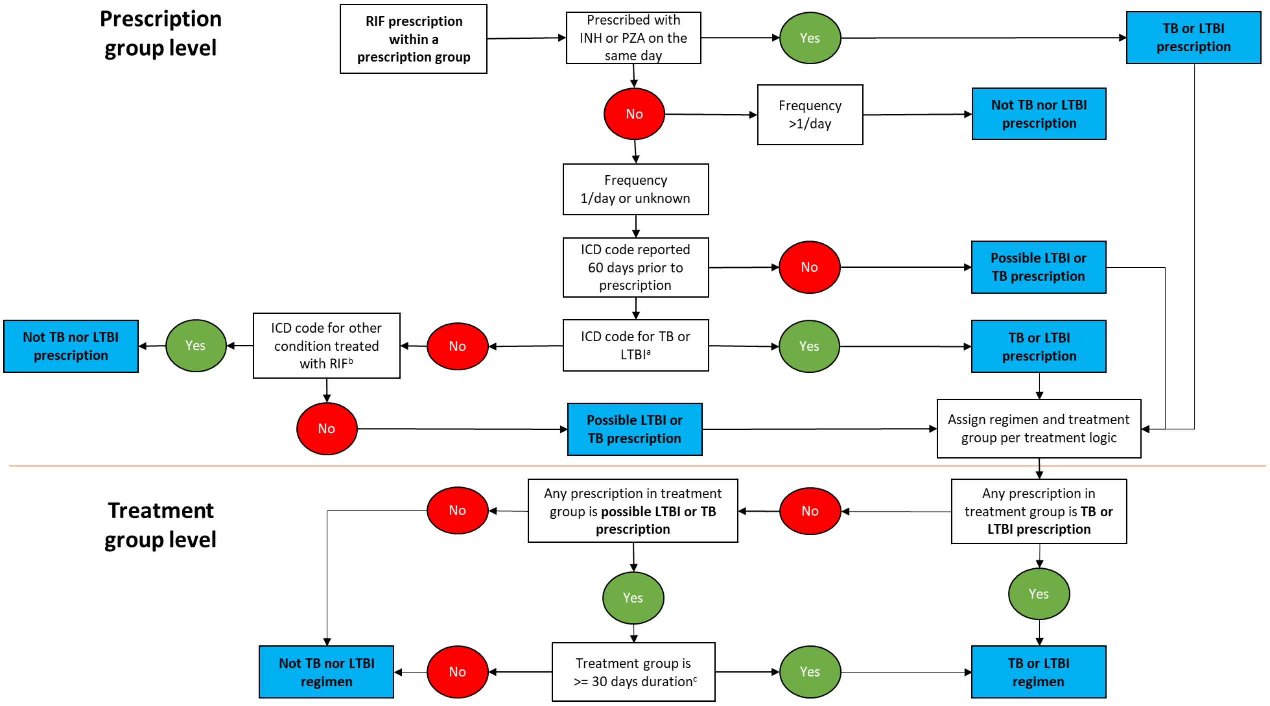


[a]*ICD-9* TB diagnostic codes: 010, 011, 012, 013, 014, 015, 016, 017, and 018; *ICD-9* LTBI diagnostic codes: 795.51 and 795.52; *ICD-10* TB diagnostic codes: A15.x, A17.x, A18.x, and A19.x; *ICD-10* LTBI diagnostic codes: Z22.7, R76.11, and R76.12.

[b]Anaplasmosis, Bartonella infections, brucellosis, cholestatic pruritis, Staphylococcus endocarditis, hidradenitis suppurativa, leprosy, meningococcal prophylaxis, pulmonary nontuberculous mycobacterial infections (severe and nonsevere), Staphylococcus infections, and Group A streptococcus carriage (*ICD-10* diagnostic codes in eTable 3).

[c]When prescription grouping for rifampin prescription is determined to be >30 days duration after treatment logic is applied and maximum time for treatment completion has been surpassed, then prescriptions are determined to be for TB or LTBI. When prescription grouping is determined to be ≤30 days duration and the maximum time for treatment completion has been surpassed, the prescription is determined not to be for TB or LTBI.

*ICD-9* indicates *International Classification of Diseases*, *Ninth Revision*; *ICD-10*, *International Classification of Diseases, Tenth Revision;* INH, isoniazid; LTBI, latent tuberculosis infection; PZA, pyrazinamide; RIF, rifampin or rifabutin; and TB, tuberculosis.

**eTable 3.** ***International Classification of Diseases, Tenth Revision*** **(*ICD-10*) Diagnostic Codes for Nontuberculosis Conditions Treated with Rifampin or Rifabutin**

| **Indication** | ***ICD-10* diagnostic codes** |
|---|---|
| Anaplasmosis | A79.82 |
| Bartonella infections | A44.x and A79.0 |
| Brucellosis | A23.x |
| Cholestatic pruritis | L29.8, L29.9 |
| Staphylococcus endocarditis | I33.0, T82.6x, and T82.7x |
| Hidradenitis suppurativa | L73.2 |
| Leprosy | A30.x |
| Meningococcal prophylaxis | Z20.811 |
| Pulmonary NTM infections, non severe | A31.x |
| Pulmonary NTM infections, severe | A31.x |
| Staphylococcus infections | B95.61, B95.62, B95.7, B95.8; M86.x; M00.0x, T84.5x, T84.6x, T84.7x; G00.3, G04.2, G06.x, G07, G08; R78.81; T85.7x; T81.4x; S91.x; L97.x; and L89.x |
| Group A Streptococcus carriage | Z22.338 |

Abbreviation: NTM, nontuberculosis mycobacteria.

**eTable 4. Antituberculosis Medications Used for Treatment Regimens**

| Medications |
|---|
| Isoniazid |
| Rifampin |
| Pyrazinamide |
| Ethambutol |
| Streptomycin |
| Rifabutin |
| Rifapentine |
| Ethionamide |
| Amikacin |
| Capreomycin |
| Ciprofloxacin |
| Levofloxacin |
| Ofloxacin |
| Moxifloxacin |
| Cycloserine |
| Para-amino salicylic acid |
| Linezolid |
| Bedaquiline |
| Delaminid |
| Clofazimine |
| Meropenem |
| Imipenem |
| Pretomanid |

### eTable 5. Electronic Health Record Data Elements and Calculated Variables Used to Determine Latent Tuberculosis Infection (LTBI) Treatment Regimen Completion Status

| LTBI regimen | Medication | Frequency[b] | Pill dosage (mg)[b] | Calculated doses/ month (ct)[c] | Calculated monthly dose (mg)[d] | Minimum regimen duration (days)[b,e] | Calculated minimum number of doses[f] | Calculated total minimum dosage (mg)[g] | Max regimen duration (months) |
|---|---|---|---|---|---|---|---|---|---|
| INH (6H/9H)[a] | INH | Daily | 300 | 30 | 9,000 | 180 | 144 | 43,200 | 12 months |
| | INH | Twice weekly | 900 | 8 | 7,200 | 180 | 42 | 37,800 | 12 months |
| RIF (4R) | RIF or rifabutin | Daily | 600 | 30 | 18,000 | 120 | 96 | 57,600 | 6 months |
| INH + RIF (3HR) | INH | Daily | 300 | 30 | 9,000 | 90 | 72 | 21,600 | 4 months |
| | RIF or rifabutin | Daily | 600 | 30 | 18,000 | 90 | 72 | 43,200 | 4 months |
| INH + RPT (3HP) | INH | Once weekly | 900 | 4 | 3,600 | 90 | 10 | 9,000 | 4 months |
| | RPT | Once weekly | 300–900 | 4 | 1,200–3,600 | 90 | 10 | 3,000–9,000 | 4 months |

Abbreviations: ct=count; INH, isoniazid; mg=milligram; RIF, rifampin or rifabutin; RPT, rifapentin.

[a]All calculations based on 6H regimen due to data to differentiate between 6H and 9H regimens being unavailable.

[b]Guidelines for the Treatment of Latent Tuberculosis Infection: Recommendations from the National Tuberculosis Controllers Association and CDC, 2020.[4]

[c]Calculated using medication frequency and a 30-day month.

[d]Calculated by multiplying pill dose by the calculated number of doses per month.

[e]Based on a 30-day month.

[f]Minimum number of doses to define completion of treatment was derived by multiplying the total doses for an adult by 80%.

[g]Calculated by multiplying the maximum adult dosage for a single pill dose by the minimum number of doses.

**eFigure 3. Reasons for Ineligibility Among Individuals Not Eligible for the Baseline Study Period Latent Tuberculosis Infection Care Cascade (N = 169,797)**

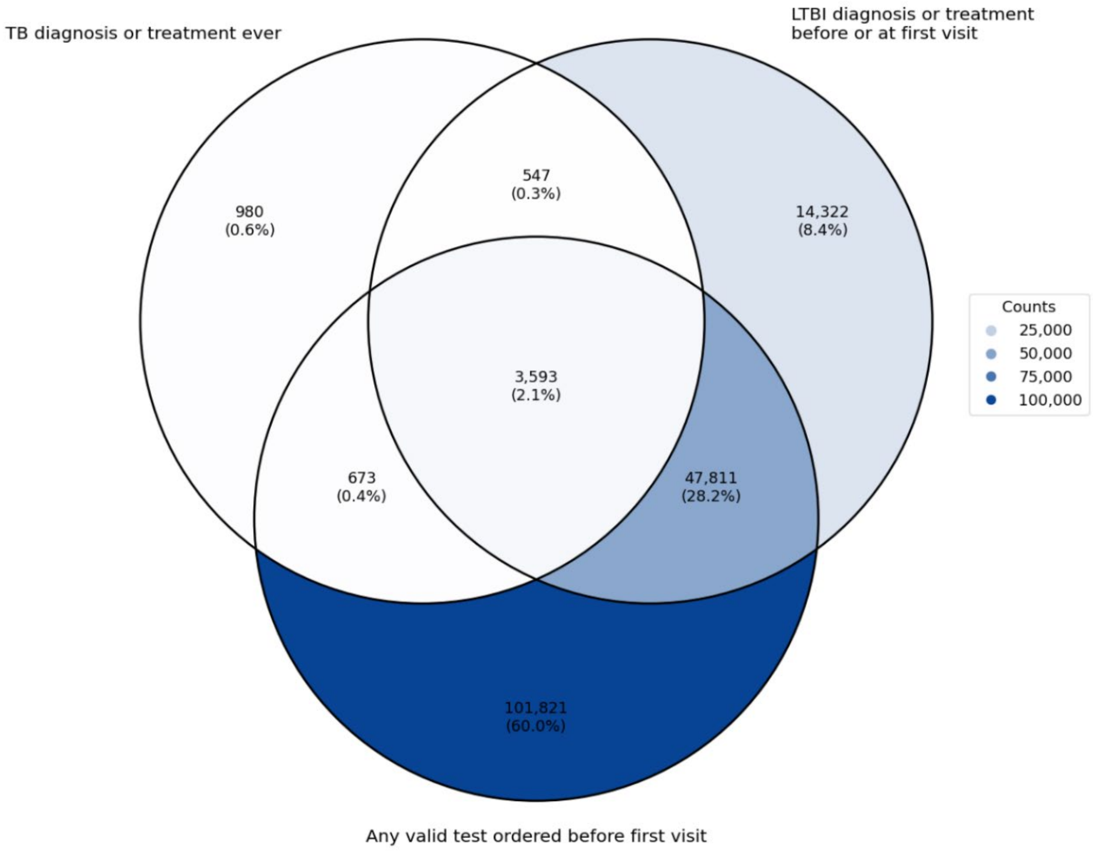


LTBI indicates latent tuberculosis infection; TB, tuberculosis disease.

**Supplemental References**


1. Centers for Disease Control and Prevention . Tuberculosis Epidemiologic Studies Consortium. Published 2024. Accessed June 12, 2024. https://www.cdc.gov/tb/research/tbesc.html

2. Prescribers' Digital Reference (PDR): Rifampin-Drug Summary. Published 2022. https://www.pdr.net/drug-summary/Rifadin-rifampin-1036

3. Vonnahme LA, Raykin J, Jones M, et al. Using electronic health record data to measure the latent tuberculosis infection care cascade in safety-net primary care clinics. *AJPM Focus*. 2023;2(4):100148. doi:https://doi.org/10.1016/j.focus.2023.100148

4. Sterling TR, Njie G, Zenner D, et al. Guidelines for the treatment of latent tuberculosis infection: recommendations from the National Tuberculosis Controllers Association and CDC, 2020. *MMWR Morb Mortal Wkly Rep*. Published online 2020. doi:10.1111/ajt.15841

5. Horsburgh CR, Goldberg S, Bethel J, et al. Latent TB infection treatment acceptance and completion in the United States and Canada. *Chest*. 2010;137(2):401-409. doi:10.1378/chest.09-0394

6. Al-Darraji HAA, Kamarulzaman A, Altice FL. Isoniazid preventive therapy in correctional facilities: a systematic review. *Int J Tuberc Lung Dis*. 2012;16(7):871-879. doi:10.5588/ijtld.11.0447

7. Sharma SK, Sharma A, Kadhiravan T, Tharyan P. Rifamycins (rifampicin, rifabutin and rifapentine) compared to isoniazid for preventing tuberculosis in HIV-negative people at risk of active TB. *Cochrane Database Syst Rev*. 2013;2013(7):CD007545. doi:10.1002/14651858.CD007545.pub2

8. Pease C, Hutton B, Yazdi F, et al. Efficacy and completion rates of rifapentine and isoniazid (3HP) compared to other treatment regimens for latent tuberculosis infection: a systematic review with network meta-analyses. *BMC Infect Dis*. 2017;17(1):265. doi:10.1186/s12879-017-2377-x

9. Codecasa LR, Murgia N, Ferrarese M, et al. Isoniazid preventive treatment: predictors of adverse events and treatment completion. *Int J Tuberc Lung Dis*. 2013;17(7):903-908. doi:10.5588/ijtld.12.0677

10. Hirsch-Moverman Y, Shrestha-Kuwahara R, Bethel J, et al. Latent tuberculous infection in the United States and Canada: who completes treatment and why? *Int J Tuberc Lung Dis*. 2015;19(1):31-38. doi:10.5588/ijtld.14.0373

11. Hall RG, Leff RD, Gumbo T. Treatment of active pulmonary tuberculosis in adults: current standards and recent advances. Insights from the Society of Infectious Diseases Pharmacists. *Pharmacotherapy*. 2009;29(12):1468-1481. doi:10.1592/phco.29.12.1468
